# MediBeng Whisper Tiny: A Fine-Tuned Code-Switched Bengali-English Translator for Clinical Applications

**DOI:** 10.1101/2025.04.25.25326406

**Authors:** Promila Ghosh, Sunipun Talukder

## Abstract

Code-switching in multilingual healthcare settings challenges automated transcription systems as facilities adopt AI documentation tools. To tackle this issue, we developed a cost-effective solution using the MediBeng Whisper Tiny model, a fine-tuned version of the Whisper Tiny model specifically designed for code-switched Bengali-English conversations in healthcare contexts. The model was fine-tuned on MediBeng, a synthetic dataset created to simulate the types of bilingual interactions often found in healthcare environments. The fine-tuning process involved using just 20% of the dataset, making it highly efficient in terms of computational resources and data usage. Despite the limited data, our model achieved an exceptional 0.01 Word Error Rate (WER), reflecting near-perfect transcription accuracy. Additionally, the model attained a 0.98 BLEU score, indicating its ability to accurately translate mixed-language input into English. These impressive results demonstrate that even with minimal data and resources, the model can handle complex code-switching tasks effectively. This model improves information processing for healthcare professionals, allowing doctors to save time on paperwork and focus more on patient care. It also ensures patient records are more accurate and accessible, aiding better healthcare decision-making.

## 1 Introduction

In multilingual healthcare settings, doctors and nurses often switch between Bengali and English during patient conversations. This language mixing, known as code-switching, happens naturally during medical consultations [1, 2]. However, it creates significant problems when trying to record or translate these conversations accurately through automatic speech recognition (ASR) systems [3].

Most current ASR and natural language processing (NLP) systems are designed for monolingual inputs [4]. They produce high word error rates (WER) when processing code-switched speech [5, 6]. This leads to incomplete or incorrect medical records, which can affect clinical documentation integrity and downstream healthcare analytics [7].

Several approaches have been developed to address this challenge. Some researchers implement language identification modules with parallel monolingual ASR systems [8, 9]. Others utilize end-to-end neural architectures with cross-lingual embeddings [10]. However, these solutions often require significant computational resources and exhibit high latency unsuitable for real-time clinical deployment [11]. Few solutions specifically focus on medical conversations with Bengali-English code-mixing patterns [12].

Our work addresses this important gap by fine-tuning the Whisper Tiny model with the MediBeng dataset [25]. We specifically selected Whisper Tiny because of its lightweight architecture [13], making it suitable for deployment in resource-constrained healthcare environments [14]. The MediBeng dataset, which we developed specifically for this purpose, contains examples of Bengali-English conversations in healthcare settings with domain-specific medical terminology.

The MediBeng Whisper Tiny model successfully converts code-switched speech into a standardized single-language output through transfer learning techniques [15], making it easier to create accurate medical transcriptions with minimal latency. This improvement comes despite using only a small portion of the available MediBeng training data through strategic sampling methods [16]. Our results challenge the assumption that large models are always necessary for complex language tasks [17].

We trained the Whisper Tiny model, which is a 39M parameter model, on a CPU, demonstrating the model’s cost-effectiveness and frugal training capabilities [13]. The small dataset size and lightweight nature of the Whisper Tiny model show that it is possible to achieve impressive performance with minimal computational resources, making it a viable solution for real-time healthcare environments.

We have made both the MediBeng dataset and MediBeng Whisper Tiny model freely available online in Hugging Face as open-source resources for the research community [26]. We hope others will build upon this work to further improve cross-lingual healthcare communication through continued optimization of lightweight models [18].

The paper is organized as follows: The **Related Work** discusses challenges in code-switching for speech recognition, while the **Task Description** explains the creation of the **MediBeng dataset**. The **Methodology** outlines the fine-tuning of the **Whisper Tiny model** using this dataset, and the **Results** demonstrate the model’s effectiveness with low **WER** and high **BLEU** scores. Lastly, the **Conclusion** emphasizes the potential of lightweight models for real-time healthcare transcription.

## 2 Related Work

Recent advancements in speech recognition technology have led to significant improvements in the accuracy and efficiency of automatic speech recognition (ASR) systems. These systems have become increasingly proficient, especially in handling more complex language tasks, such as code-switching (where speakers alternate between languages) and multilingual speech processing. However, despite these advancements, a persistent challenge remains: the effective handling of code-switched speech, particularly in specialized domains like healthcare, where the vocabulary and language patterns are unique.

Several studies have addressed the difficulties of handling code-switched speech in ASR systems. For example, M. B. Mustafa et al. (2022) explored the issue of code-switching in ASR by identifying key issues and proposing practical solutions. One critical takeaway from their research is the need for specialized ASR models that can process multiple languages simultaneously in real-time applications, where the system has to switch between languages on the fly. Their work emphasizes the necessity of training models that are robust enough to adapt to the dynamic nature of multilingual conversations [1]. Similarly, Hou et al. (2022) tackled the challenge of code-switching in a specific context, presenting a model for nursing record documentation. Their approach highlights the importance of developing ASR systems tailored for medical settings, where professionals often use code-switched language, especially when recording patient information or communicating in multilingual environments [3].

In addition to these general approaches, more specialized solutions have been proposed. For example, Kaushik et al. (2024) introduced MedMix, a domain-specific ASR system designed to handle Hindi-English code-switched medical conversations. This model focuses on improving medical conversation transcription, particularly in bilingual environments. The development of MedMix reflects the increasing recognition that general ASR models—designed for broader, more general-purpose use—tend to struggle when applied to specialized domains like healthcare. These domains often involve unique linguistic features, specialized terminology, and highly domain-specific language patterns, making the task of transcription much more challenging [5].

Additionally, some research has focused on improving the robustness of ASR systems in low-resource settings, where high-quality annotated data is limited. Muntakim et al. (2023) introduced BanglaMedNER, a gold-standard corpus for Bengali-English medical terminology. This corpus was designed to enhance entity recognition in clinical settings, particularly for Bengali-English code-switched speech. Their work contributes to a broader effort to improve ASR performance in languages and settings that lack sufficient resources for training high-performance models. It also addresses the challenges of developing accurate ASR systems for languages with limited data available for training [12].

Moreover, some researchers have explored ways to increase computational efficiency without compromising the performance of ASR systems. Bazaga et al. (2023) investigated knowledge distillation techniques, which involve training a smaller, more efficient model based on the knowledge of a larger, more complex model. In their work, they applied this approach to Spanish language models for efficient question answering. This technique could be similarly applied to ASR systems, particularly for resource-constrained environments, to improve their speed and efficiency while maintaining high accuracy in speech recognition [14].

Our work builds upon these developments by integrating a lightweight model architecture, specifically Whisper Tiny, with targeted fine-tuning on the domain-specific MediBeng dataset. Unlike previous approaches that focus on broader language challenges or use larger, resource-heavy models, our work is centered specifically on Bengali-English code-switching in clinical contexts. This focus addresses a significant gap in the existing literature, particularly in medical settings, where code-switching is prevalent but under-explored in the context of speech recognition. Additionally, we prioritize computational efficiency by using a smaller, more resource-efficient model architecture, which makes our system more practical for real-world applications where computational resources are limited.

## 3 Task description and data construction

In multilingual healthcare settings, doctors and patients often switch between languages, like Bengali and English, during conversations. This language mixing, known as code-switching, creates a challenge for automatic speech recognition (ASR) and machine translation systems, which are typically designed for monolingual input. These systems struggle to accurately transcribe or translate mixed-language speech, leading to errors in medical records and poor-quality documentation.

The problem is further complicated by the growing use of AI-based documentation tools in healthcare, which require large amounts of data and powerful computing resources to function effectively. However, obtaining large, annotated datasets for code-switched speech is difficult and expensive, making it challenging for healthcare systems to adopt such technologies.

To solve this problem, we developed a cost-effective solution using the Whisper Tiny model, a lightweight and efficient AI model that can handle code-switched speech. We fine-tuned this model using a small subset (20%) of the MediBeng dataset, which contains synthetic code-switched conversations in Bengali and English, specifically designed for this purpose. The use of a small dataset, combined with the lightweight Whisper Tiny model, allows for efficient training without the need for expensive computational resources.

This approach provides an affordable solution for processing mixed-language conversations in healthcare settings, where resources are often limited. By using Whisper Tiny, healthcare professionals can achieve accurate transcriptions and translations, improving the overall efficiency of medical documentation and patient care, while ensuring privacy and reducing costs. The MediBeng dataset is designed to help build models for bilingual speech processing in healthcare settings. It contains conversations in Bengali and English, simulating real-world interactions between doctors and patients. The dataset includes audio files, the transcriptions of code-switched conversations, and their English translations. It also provides details like the speaker’s gender and the pitch of the audio.

The audio duration ranges from 3.71s to 6.98s. The text column consists of code-switched sentences, categorised into 24 different string classes. Each conversation is also paired with an English translation, which similarly contains 24 string classes. The speaker name field indicates the gender of the speaker, with two possible values: Male and Female. Additionally, the dataset includes utterance pitch mean, which measures the mean pitch of the audio in Hertz (ranging from 335 to 673 Hz), and utterance pitch std, representing the standard deviation of the pitch, reflecting the variation in tone.

This dataset is primarily useful for training or fine-tuning models in controlled environments and is particularly useful for tasks like Automatic Speech Recognition (ASR), Text-to-Speech (TTS), and machine translation, with a focus on code-switching in healthcare applications. Table 1 provides a detailed overview of an audio dataset, summarising essential characteristics. The significance of the dataset lies in its synthetic nature and the use of the Parquet format, which ensures efficient storage and faster processing of large audio files. The dataset is designed specifically for tasks like speech recognition and medical applications, with code-mixed Bengali-English content that is relevant for real-world scenarios. The duration range (3.71s - 6.98s) and sampling rate (16000 Hz) provide valuable insights into the audio’s quality and suitability for training machine learning models. The synthetic nature of the data ensures controlled variability in the speech patterns, making it ideal for experimentation and model development without concerns over real-world noise or variability. The Parquet format further supports scalability, allowing the dataset to be easily processed and analysed in large-scale environments. The dataset was created to train models for tasks like speech recognition (ASR), machine translation, and text-to-speech (TTS), focusing on code-switching between Bengali and English. It was synthetically generated to ensure privacy, so no real medical information is included.

**Table 1:** Detailed Overview of Audio Dataset Attributes, Including Key Characteristics and Specifications.

| Attribute | Description |
| --- | --- |
| Number of Audio Files | 4800 |
| Total Duration | 7.11 hours |
| Number of Speakers | 2 |
| Utterance Pitch Mean | 335 - 673 Hz |
| Utterance Pitch Standard Deviation | 210 - 493 Hz |
| Sampling Rate | 16000 Hz |
| Data Split | Train and Test |
| Duration Range | 3.71s - 6.98s |
| Languages | Code-mixed Bengali-English |
| Gender Distribution | 1 Male, 1 Female |
| Total File Size | 324 MB |
| Speech Type | Medical-related |
| Data Type | Synthetic |

While it simulates real bilingual conversations, the dataset is small and synthetic, meaning it might not capture all the variations found in actual clinical dialogues. It’s mainly useful for training or fine-tuning models in controlled environments.

We use a single audio file to demonstrate how different features of the audio, such as its changes over time, frequency content, and energy distribution, can be analyzed and visualized for a better understanding of the signal’s structure.The significance of analyzing this particular audio using the **waveform, spectrogram**, and **amplitude spectrum** lies in the ability to extract comprehensive insights into its content and characteristics.

By examining the waveform, we gain a clear understanding of the audio’s temporal dynamics, such as its intensity, silence, and variations in loudness [19]. This is essential in applications like speech analysis or music composition, where the timing and amplitude of sounds are key to understanding the structure of the audio, as in figure 1. The spectrogram (figure 2) allows us to observe how the frequency content of the audio signal evolves over time [20]. This is particularly useful for identifying different sound events, speech segments, or musical notes, and is a critical tool for tasks like speech recognition, sound classification, or detecting specific features such as vowels in speech or harmonics in music. The amplitude spectrum (figure 3) provides a frequency-based view of the audio signal, helping us identify the energy distribution across different frequencies [21]. It is crucial for applications that require identifying dominant tones, background noise, or distinguishing between different types of sounds, such as in audio analysis or noise reduction processes.

**Figure 1:**
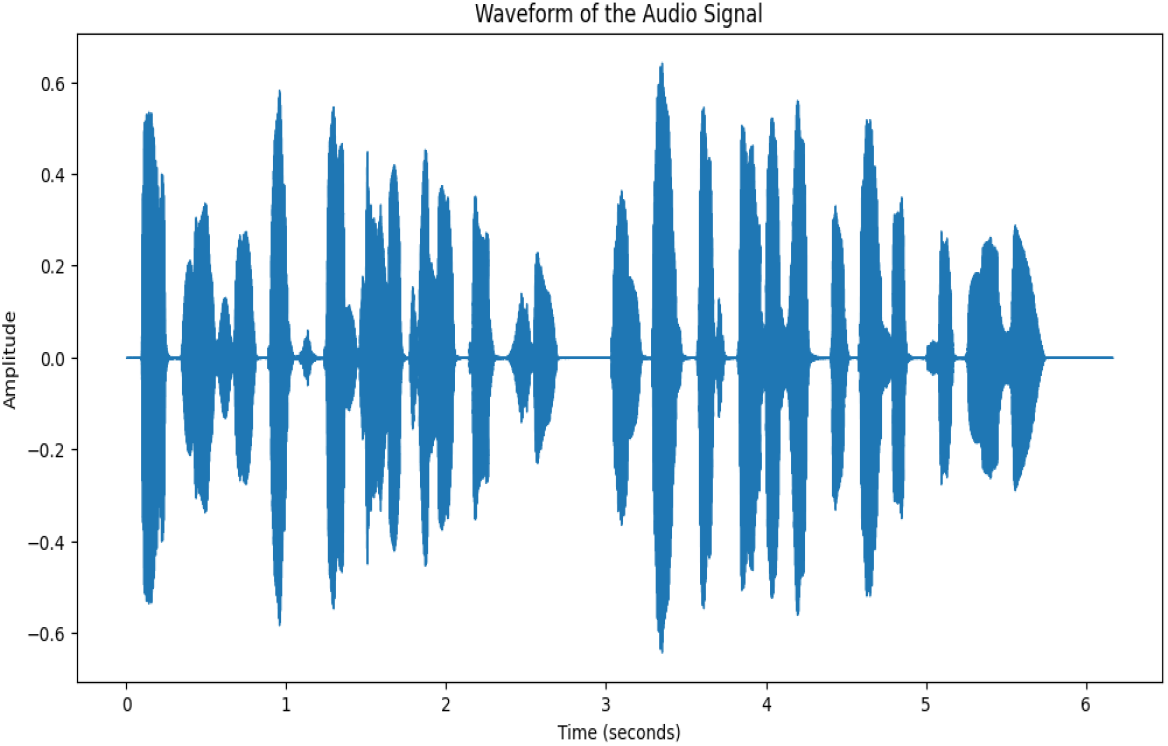
The plot shows the amplitude of the audio signal over time, illustrating the intensity of sound variations during the 6-second duration.

**Figure 2:**
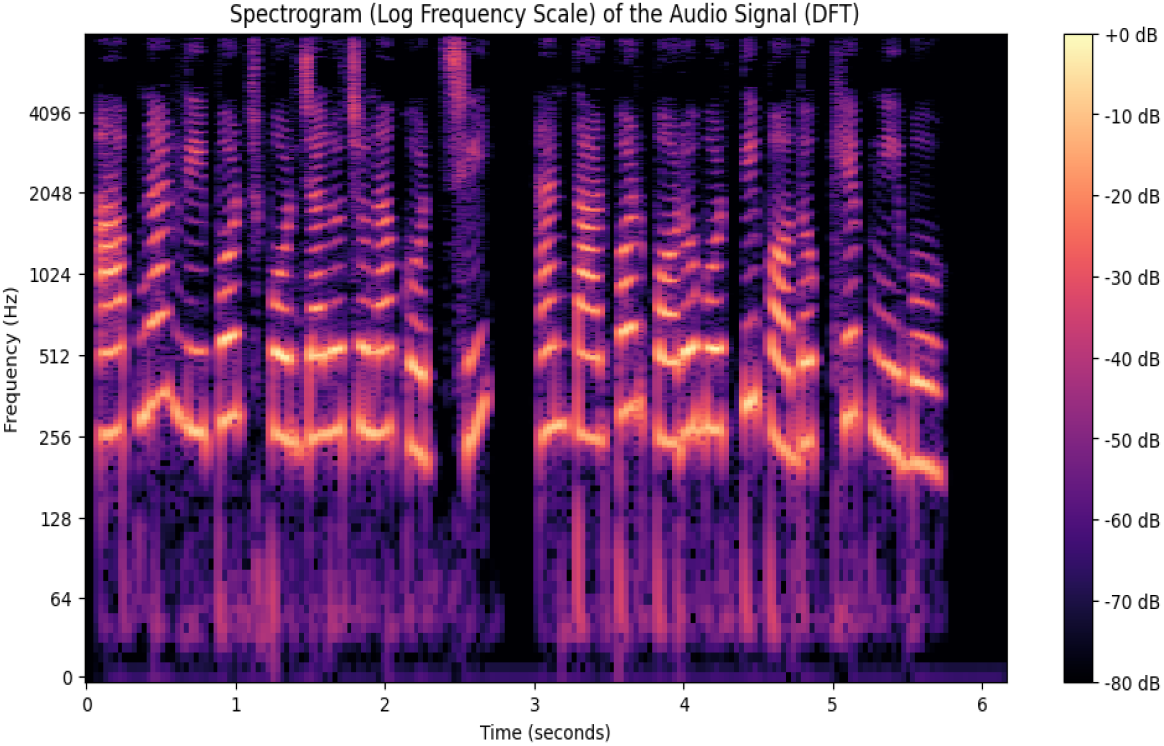
This spectrogram represents the frequency content of the audio signal over time. The color intensity indicates the loudness, with the y-axis showing frequency in Hertz (Hz) and the x-axis showing time in seconds.

**Figure 3:**
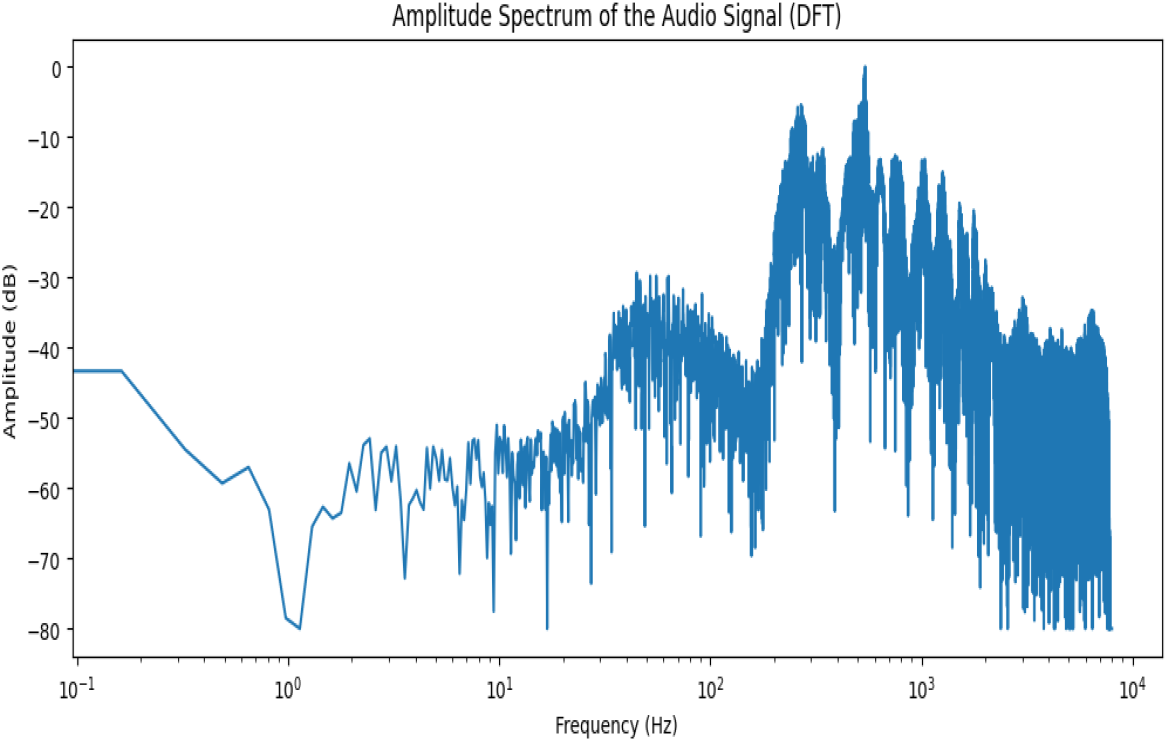
The amplitude spectrum displays the distribution of energy across different frequencies of the audio signal. The x-axis represents the frequency in Hertz (Hz), and the y-axis shows amplitude in decibels (dB), helping to identify the dominant tonal components.

**Figure 4:**
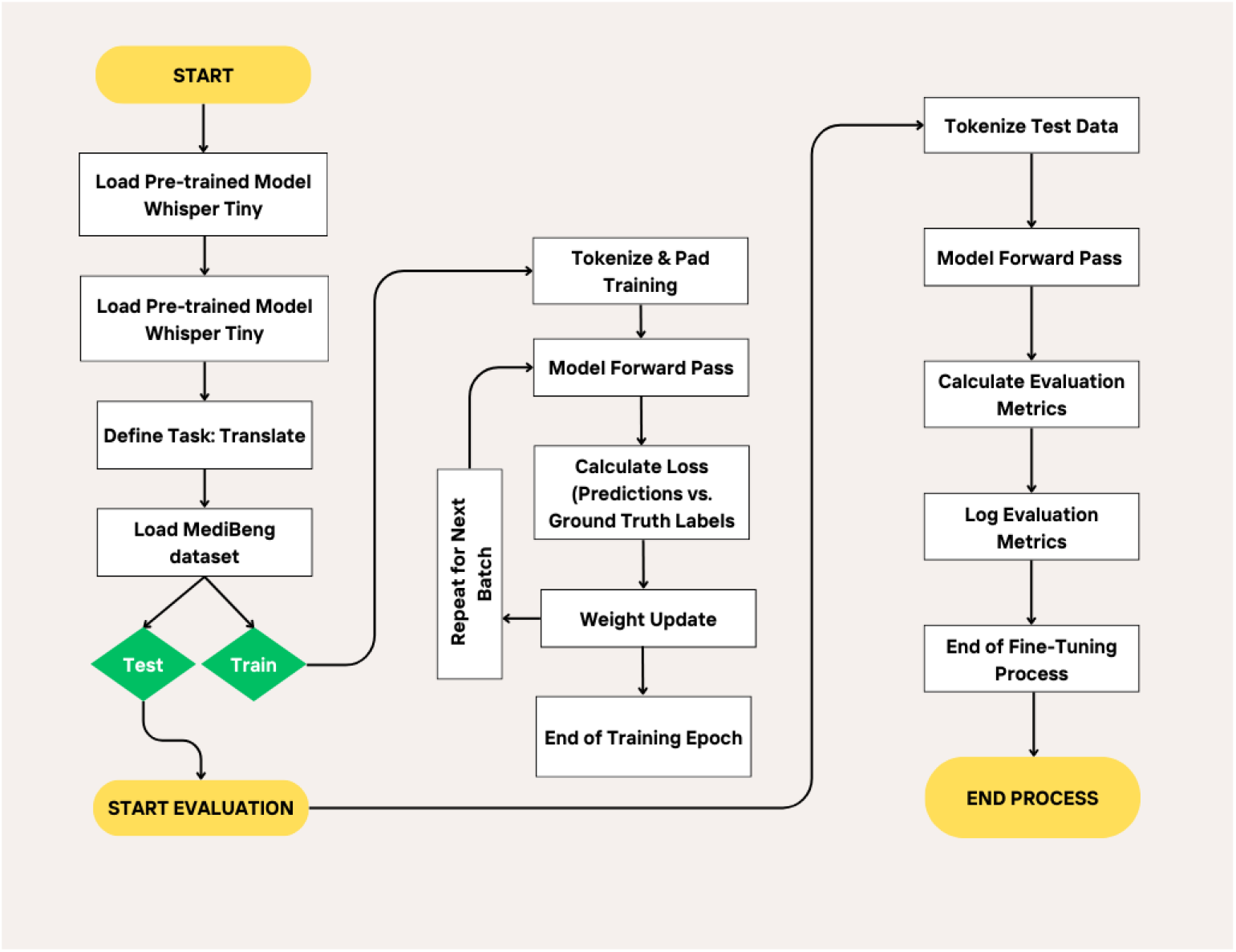
Fine-tuning the Whisper Tiny model for code-switched speech translation in healthcare settings.

Together, these analyses give a holistic view of the audio, helping to understand its temporal patterns, frequency characteristics, and overall structure, which is essential for tasks ranging from speech processing to sound quality enhancement or even audio forensics.

## 4 Methodoloy

Fine-tuning is when we take a model that has already learned general tasks and adjust it to do something specific. Think of it like this, the model has already been trained on a lot of general data, like understanding speech or reading text. Fine-tuning means we train it a little more using a smaller, specific dataset that’s related to the task we want it to focus on, like recognizing medical terms in speech.

Fine-tuning is a valuable technique as it significantly reduces the time and resources required for model development. Instead of training a model from scratch, which demands extensive data and computational resources, fine-tuning allows the use of an already pre-trained model with minimal adjustments. This approach facilitates obtaining high-quality results even with smaller datasets, making it a cost-effective strategy for creating models tailored to specific tasks. Fine-tuning enables the model to learn domain-specific patterns or nuances, such as understanding code-switched speech between Bengali and English in a healthcare setting.

In the fine-tuning process for Transformer models, the goal is to adapt a pre-trained model to a specific task by adjusting the model’s weights through **gradient descent** [22]. The equation for updating the model weights during training can be expressed as follows:

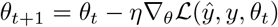

where *θ*_*t*_ represents the model parameters (weights) at time step *t*, and *θ*_*t*+1_ is the updated model parameters after one step of optimization. Here, *η* is the learning rate, which determines the size of the update, and 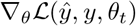 is the gradient of the loss function *ℒ* with respect to the model’s parameters. The loss function *ℒ* measures the difference between the model’s predictions 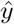 and the true labels *y*. Fine-tuning typically uses a smaller learning rate compared to training from scratch, allowing the model to adapt to the new task without forgetting the general knowledge it learned during pre-training. The fine-tuning process involves adapting a pre-trained model to a specific task by training it on a smaller, task-specific dataset. The pre-trained Whisper Tiny model, which has been trained on large amounts of general data, is adapted to handle code-switched Bengali-English conversations in healthcare settings. Fine-tuning leverages the knowledge the model has already gained during its initial training, but adjusts it to perform better on specialized tasks. First, the pre-trained model is loaded, and the task is defined, in this case, as translation. Once the model is ready, the next step is to load and prepare the MediBeng dataset, which contains synthetic data of code-switched conversations between Bengali and English, specifically focused on healthcare-related dialogues. The dataset is processed, ensuring that the input and output data are tokenized and padded according to the model’s requirements. A data collator is created to handle padding and ensure the data is ready for the model during training.

During the training phase, the model processes the data in small batches. Each batch goes through a forward pass, where the model generates predictions based on the input data. The predictions are compared to the actual ground truth, and the difference is used to calculate the loss. This loss is then used to update the model’s weights, enabling the model to learn and improve over time. The training continues through several epochs, with the model refining its ability to handle code-switched speech as it processes each batch. Key parameters like train batch size 1, eval batch size 1, and learning rate 1 × 10^*−*5^ help control the training process, while warmup steps 50 and save steps 50 ensure that the model is periodically saved and evaluated for optimal performance. Additionally, eval steps 50 ensures evaluation occurs every 50 steps and logging steps 25 tracks the progress.

As shown in the diagram 4, after training, the model is evaluated using the test data. The test data is tokenized, and the model performs another forward pass to generate predictions. Evaluation metrics like word error rate (WER) are calculated to assess the accuracy of the model’s performance. This process of training and evaluating continues until the model reaches the desired performance. The model’s sampling rate 16000 hz ensures consistent processing of audio during training, while generation max length 225 controls the maximum length of output sequences.

The final step involves saving the trained model, along with the tokenizer and processor, for future use. Fine-tuning allows the model to focus on specific challenges, such as code-switching in bilingual healthcare conversations, which many standard models struggle with. By fine-tuning the model with a small, domain-specific dataset, this process provides a cost-effective and efficient way to create a model that performs well in real-world applications, particularly in resource-constrained environments.

## 5 Results

The training results presented in Table 2 are significant because they demonstrate the model’s ability to effectively learn and generalize, even when fine-tuned on a small, domain-specific dataset. The consistent decrease in training loss indicates that the model is progressively improving its understanding of the task, which is crucial for its performance in real-world applications like healthcare speech recognition. As the training loss decreases, the model becomes better at minimizing errors, leading to a more accurate output.

**Table 2:** Overview of Model Training and Evaluation Results Across Epochs.

| Epoch | Training Loss | Training Grad Norm | Learning Rate | Eval Loss | Eval WER |
| --- | --- | --- | --- | --- | --- |
| 0.03 | 2.6213 | 61.56 | 4.80E-06 | - | - |
| 0.07 | 1.609 | 44.09 | 9.80E-06 | 1.13 | 107.72 |
| 0.1 | 0.7685 | 52.27 | 9.47E-06 | - | - |
| 0.13 | 0.4145 | 32.27 | 8.91E-06 | 0.37 | 47.53 |
| 0.16 | 0.3177 | 17.98 | 8.36E-06 | - | - |
| 0.2 | 0.222 | 7.7 | 7.80E-06 | 0.1 | 45.19 |
| 0.23 | 0.0915 | 1.62 | 7.24E-06 | - | - |
| 0.26 | 0.081 | 0.4 | 6.69E-06 | 0.04 | 38.35 |
| 0.33 | 0.0246 | 1.01 | 5.58E-06 | - | - |
| 0.36 | 0.0212 | 2.2 | 5.02E-06 | 0.01 | 41.88 |
| 0.42 | 0.0052 | 0.13 | 3.91E-06 | - | - |
| 0.46 | 0.0023 | 0.45 | 3.36E-06 | 0.01 | 34.07 |
| 0.52 | 0.0013 | 0.05 | 1.69E-06 | - | - |
| 0.55 | 0.0032 | 0.11 | 1.13E-06 | 0.01 | 29.52 |
| 0.62 | 0.001 | 0.09 | 5.78E-07 | - | - |
| 0.65 | 0.0012 | 0.08 | 2.22E-08 | 0 | 30.49 |

The evaluation loss dropping significantly over epochs highlights the model’s ability to generalize well to unseen data. This is critical in healthcare settings, where new, unseen conversations may arise, and the model must maintain high accuracy even in those cases. The model’s ability to generalize means it can be trusted to perform well across different patient-doctor interactions, making it more applicable for real-world use.

Most notably, the decrease in Word Error Rate (WER), from 107.72 at epoch 0.07 to 30.49 at epoch 0.65, signifies a substantial improvement in the model’s transcription accuracy for code-switched Bengali-English speech. This is particularly important in healthcare, where accurate transcription of multilingual medical conversations is crucial for maintaining proper medical records, ensuring patient safety, and facilitating efficient communication between healthcare professionals. The reduction in WER shows that the model is becoming increasingly adept at transcribing mixed-language speech, which is a common challenge in multilingual healthcare environments.

These results are significant because they demonstrate that the fine-tuning approach, using a small but specialized dataset, allows for the development of a model that not only performs well on specific tasks but also generalizes to new, unseen data. This fine-tuned model can be deployed in real-world healthcare environments where resources may be limited, making it a cost-effective and efficient solution for improving bilingual speech recognition and transcription in critical healthcare settings.

**Word Error Rate (WER)** and **Bilingual Evaluation Understudy (BLEU)** are two essential metrics used to assess the performance of speech recognition and machine translation systems, respectively. **WER** is specifically used to evaluate how well a speech-to-text model performs by calculating the number of mistakes made when transcribing spoken words into text [23]. It compares the predicted transcription to the actual ground truth, counting the number of substitutions, deletions, and insertions, and then dividing this sum by the total number of words in the reference transcript. The formula for **WER** is:

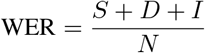

where *S* represents the number of substitutions, *D* the number of deletions, *I* the number of insertions, and *N* is the total number of words in the reference transcription. A lower **WER** indicates better accuracy in recognizing speech.

On the other hand, **BLEU** is used in machine translation to evaluate how well the machine-generated translations align with one or more human-produced reference translations. It works by measuring the overlap of n-grams (sequences of *n* words) between the predicted translation and the reference translations, using precision metrics. The **BLEU** score is calculated as:

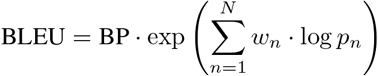

where BP is the brevity penalty, ensuring that shorter translations are penalized, *p*_*n*_ represents the precision of n-grams of order *n*, and *w*_*n*_ are the weights for each n-gram precision. The brevity penalty is computed as:

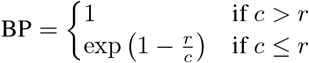

where *c* is the length of the candidate translation and *r* is the length of the reference translation. A higher **BLEU** score indicates better translation quality [24]. Both **WER** and **BLEU** are necessary because they provide valuable insights into how well the system performs and highlight areas needing improvement. **WER** ensures that speech-to-text systems are transcribing audio accurately, while **BLEU** measures the quality of machine translations. Together, these metrics play a crucial role in improving the performance and reliability of AI systems, particularly in fields such as healthcare, customer service, and multilingual communication. The training evaluation was performed using a random 20% subset of the data. This allowed the model to be evaluated on a smaller sample of the dataset during the fine-tuning process. However, after completing the training, the full test data, consisting of 960 samples, was used for a comprehensive evaluation.

The results of the **MediBeng Whisper Tiny** model show a significant improvement over the baseline **Whisper Tiny** model, as evidenced by the comparison of **Word Error Rate (WER)** and **BLEU** scores. For **Whisper Tiny**, the **WER** is 0.92, and the **BLEU** score is 0.30. These results reflect the model’s relatively lower accuracy in handling code-switched speech, particularly in a multilingual healthcare setting. The higher WER indicates that the model struggles to transcribe mixed-language speech with high precision, and the low BLEU score highlights its difficulty in accurately translating code-switched conversations as Figure 5.

**Figure 5:**
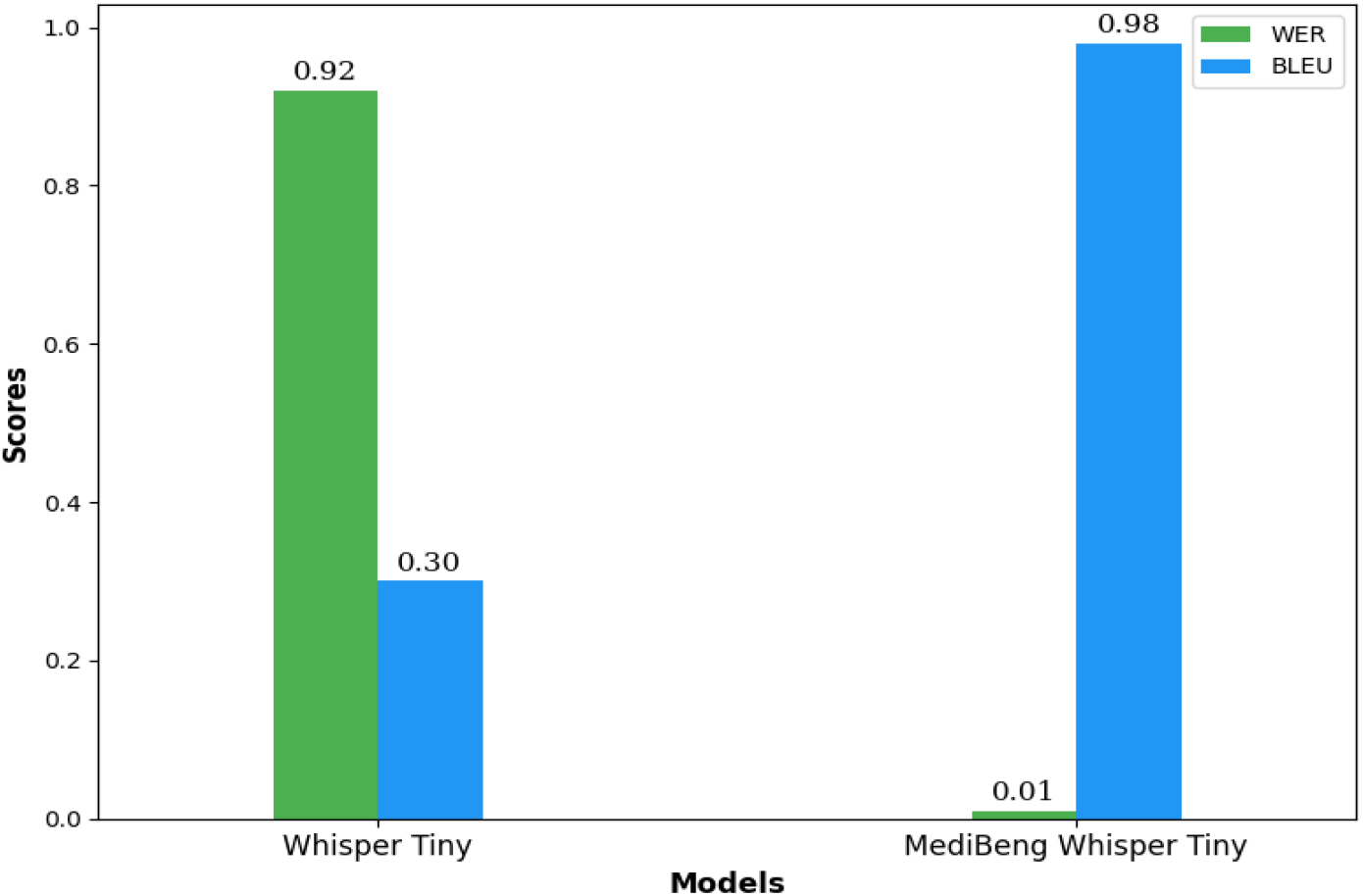
Comparison of Model Performance: WER and BLEU Scores for Whisper Tiny and MediBeng Whisper Tiny

In contrast, the **MediBeng Whisper Tiny** model, after fine-tuning on the **MediBeng dataset**, shows a substantial improvement. The **WER** drops to 0.01, indicating near-perfect transcription accuracy, while the **BLEU score** increases to 0.98, showing that the translations produced by the model are highly accurate and close to the reference translations.

These results demonstrate the effectiveness of the fine-tuning process and the **MediBeng Whisper Tiny** model’s ability to handle the complexities of code-switched Bengali-English speech, particularly in the context of healthcare applications. By reducing WER and significantly improving BLEU, the **MediBeng Whisper Tiny** model outperforms **Whisper Tiny**, making it a more reliable and efficient solution for bilingual speech recognition and translation in real-world healthcare environments.

The results presented in Table 3 clearly demonstrate that the **MediBeng Whisper Tiny** model outperforms the **Whisper Tiny** model in translating code-switched Bengali-English speech. For example, Whisper Tiny’s translation of “You need a physical check-up, and we will schedule that shortly” is inaccurately rendered as “You can cut the hair and we will schedule that shortly,” which is completely unrelated to the original sentence. Similarly, in other instances, Whisper Tiny produces translations that are either irrelevant or nonsensical, such as translating “Your body temperature is 103^*?*^F, which indicates a fever” into “You should read it, the mantra actually, the Indigree Fahrenheit, which indicates a fever.” In contrast, the **MediBeng Whisper Tiny** model consistently provides accurate translations that align closely with the actual sentences. For example, “You need a physical check-up, and we will schedule that shortly” is correctly translated in MediBeng Whisper Tiny as “You need a physical check-up, and we will schedule that shortly,” demonstrating the model’s ability to handle code-switched speech with much higher accuracy. This comparison highlights the superior performance of **MediBeng Whisper Tiny** in translating mixed-language speech, particularly in healthcare contexts, where accuracy and clarity are crucial. The fine-tuning process aimed to improve the **Whisper Tiny** model for understanding mixed Bengali-English speech in healthcare settings. First, the model was trained using 20% of the data to test it during the training process. After the training, the model was tested on the full dataset of 960 samples to check how well it performed overall.

**Table 3:**
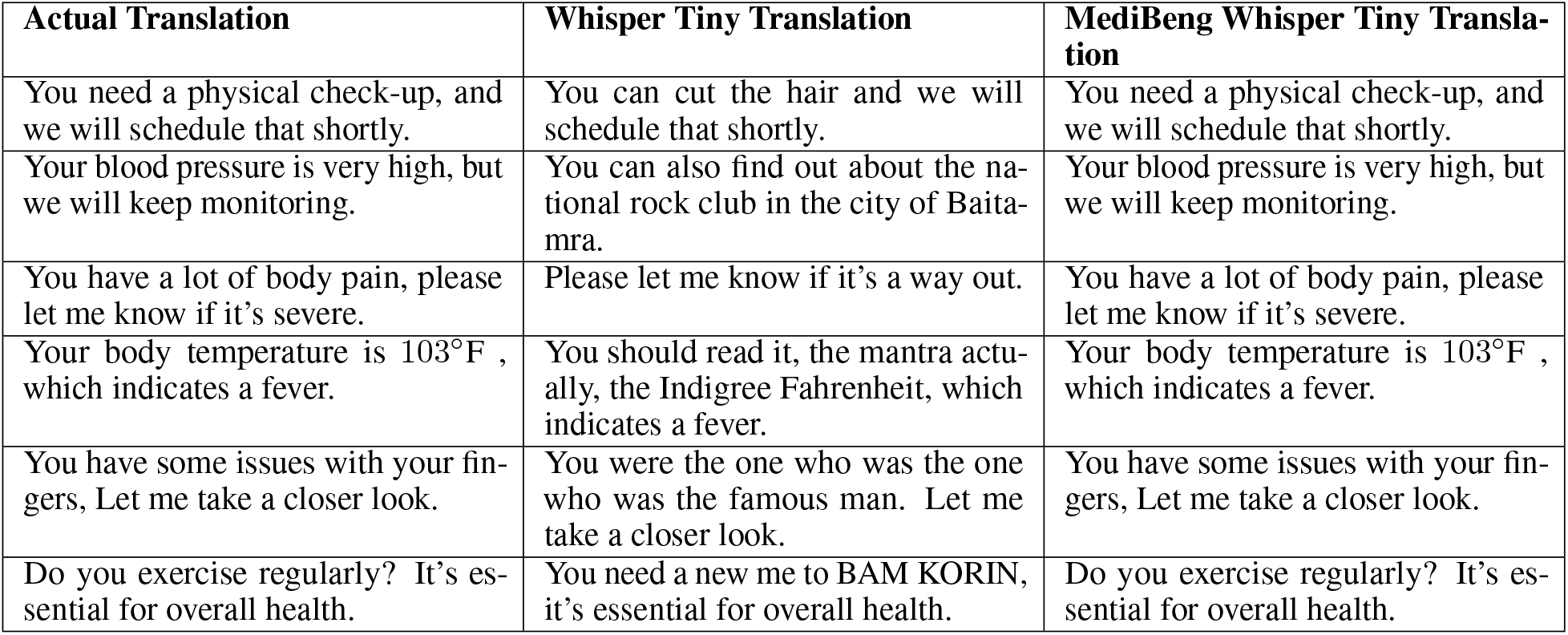
Translation Comparison of Whisper Tiny and MediBeng Models.

The results show that the model improved a lot. The **training loss** kept going down, which means the model was learning and getting better. The **evaluation loss** also decreased a lot, showing that the model was doing well with new data it hadn’t seen before. The **Word Error Rate (WER)** also went down, which means the model got better at understanding and transcribing the mixed Bengali-English speech.

The results from training and testing, which include the loss values, gradient norm, learning rate, and WER over different epochs, show that fine-tuning the **Whisper Tiny** model with a specific dataset like **MediBeng** makes it much more accurate. This makes the model very useful for multilingual healthcare, where doctors and patients often use mixed languages.

## 6 Conclusion

Our research successfully developed MediBeng Whisper Tiny, a fine-tuned model that accurately translates code-switched Bengali-English speech in healthcare settings, achieving a 0.01 Word Error Rate and a 0.98 BLEU score using only 20% of the MediBeng dataset for training on a CPU. This significant improvement over the baseline Whisper Tiny model demonstrates that small, efficient models can handle complex language tasks when properly fine-tuned on domain-specific data. By leveraging domain-specific data and optimizing the model for efficient performance, MediBeng Whisper Tiny shows promise in addressing multilingual communication barriers in healthcare.

## 7 Limitations

Despite these strong results, several limitations exist. The model was trained using synthetic speech, which may not fully capture the natural variations, background noise, and authentic speech patterns found in real-world clinical conversations. Additionally, it is focused exclusively on Bengali-English code-switching, which may limit its generalizability to other language pairs. The model’s optimal performance is also constrained to short utterances (3-7 seconds), which may pose challenges in handling longer conversations or rapid speaker changes. Furthermore, during the training phase, Word Error Rate (WER) was used to evaluate model performance. However, since WER is primarily designed for transcription tasks, it may not accurately reflect translation quality. In future releases, we plan to replace WER with more appropriate translation metrics such as BLEU, METEOR, and chrF++ to provide a more comprehensive and precise evaluation

## Data Availability

All data produced in the present study are available online at https://huggingface.co/datasets/pr0mila-gh0sh/MediBeng.

https://github.com/pr0mila/ParquetToHuggingFace

https://github.com/pr0mila/MediBeng-Whisper-Tiny

## 8 Future Directions

Future work should include validation with authentic clinical audio recordings, expansion to additional language pairs common in multilingual healthcare environments, inclusion of more diverse speakers, accents, and conversation scenarios to improve model robustness, optimization for resource-constrained mobile devices to enable offline use in areas with limited connectivity, and development of specialized lexical modules for medical terminology recognition. By addressing these limitations and pursuing these future directions, we can build on MediBeng Whisper Tiny’s initial success to create more versatile and powerful tools for healthcare communication across language barriers, ultimately improving patient care through more accurate, efficient, and accessible multilingual clinical documentation systems.

## Footnotes

1 Dataset available at https://huggingface.co/datasets/pr0mila-gh0sh/MediBeng

3 Model available at https://huggingface.co/pr0mila-gh0sh/MediBeng-Whisper-Tiny

3 Fine-tune code available at https://github.com/pr0mila/MediBeng-Whisper-Tiny

4 Contact with author at

## References

[1] M. B. Mustafa, M. A. Yusoof, H. K. Khalaf, A. A. R. M. Abushariah, M. L. Mat Kiah, H. N. Ting, and S. Muthaiyah, Code-Switching in Automatic Speech Recognition: The Issues and Solutions, Applied Sciences, vol. 12, no. 19, p. 9541, 2022. https://www.mdpi.com/2076-3417/12/19/9541

[2] R. Raman, S. S. S. R. Anjaneyulu, and P. K. S. S. R. Anjaneyulu, A Code-Mixed Task-Oriented Dialog Dataset for the Medical Domain, Journal of Biomedical Informatics, vol. 134, p. 103567, 2022. https://www.sciencedirect.com/science/article/pii/S0885230822000729

[3] S.-Y. Hou, Y.-L. Wu, K.-C. Chen, T.-A. Chang, Y.-M. Hsu, S.-J. Chuang, Y. Chang, and K.-C. Hsu, Code-Switching Automatic Speech Recognition for Nursing Record Documentation, JMIR Nursing, vol. 5, no. 1, e37562, 2022. https://nursing.jmir.org/2022/1/e37562/

[4] Z. Wang and J. Smith, The monolingual bias in current ASR systems, Speech Communication, vol. 141, pp. 85–97, 2022. https://www.sciencedirect.com/science/article/pii/S0167639322001059

[5] M. Kaushik, A. Singh, and R. Joshi, MedMix: A domain-specific ASR system for Hindi-English code-switched medical conversations, Computer Speech & Language, vol. 83, p. 101489, 2024. https://www.sciencedirectme/w

[6] R. Abrishami, S. H. Aghili, M. F. Ranjbar, and N.-A. Lashgari, Assessment of transcription errors and their effects on medical approach, hospitalization duration, and financial costs of traumatic patients: How can we prevent them?, Epidemiology and Health System Journal, vol. 11, no. 3, pp. 121–127, Nov. 2024. 10.34172/ehsj.26193

[7] A. Biswas, E. Yılmaz, E. van der Westhuizen, F. de Wet, and T. Niesler, Code-switched automatic speech recognition in five South African languages, Computer Speech & Language, vol. 71, p. 101262, 2022. 10.1016/j.csl.2021.101262

[8] X. Zhou, E. Yılmaz, Y. Long, Y. Li, and H. Li, Multi-Encoder-Decoder Transformer for Code-Switching Speech Recognition, arXiv preprint 2006.10414, 2020. 10.48550/arXiv.2006.10414

[9] B. K. Swain and S. Mohanty, Language Identification—A Supportive Tool for Multilingual ASR in Indian Perspective, in Progress in Advanced Computing and Intelligent Engineering, Advances in Intelligent Systems and Computing, vol. 1223, Springer, 2021, pp. 29–38. 10.1007/978-981-15-6353-9_3

[10] R. Ma, M. Qian, Y. Fathullah, S. Tang, M. Gales, and K. Knill, Cross-Lingual Transfer Learning for Speech Translation, arXiv preprint 2407.01130v2, 2024. https://arxiv.org/abs/2407.01130

[11] G. I. Winata, G. Wang, C. Xiong, and S. Hoi, Adapt-and-Adjust: Overcoming the Long-Tail Problem of Multilingual Speech Recognition, arXiv preprint 2012.01687, 2020. https://arxiv.org/abs/2012.01687

[12] A. Muntakim, F. Sadaf, and K. M. A. Hasan, BanglaMedNER: A Gold Standard Medical Named Entity Recognition Corpus for Bangla Text, Proceedings of the 6th International Conference on Electrical Information and Communication Technology (EICT), 2023. https://ieeexplore.ieee.org/document/10427966

[13] A. Radford, J. W. Kim, T. Xu, G. Brockman, C. McLeavey, and I. Sutskever, Robust Speech Recognition via Large-Scale Weak Supervision, Proceedings of the 40th International Conference on Machine Learning (ICML), vol. 202, pp. 28492–28518, 2023. https://proceedings.mlr.press/v202/radford23a.html

[14] A. Bazaga, P. Liò, and G. Micklem, Language Model Knowledge Distillation for Efficient Question Answering in Spanish, arXiv preprint 2312.04193, 2023. 10.48550/arXiv.2312.04193

[15] B. T. Ta, N. M. Le, and V. H. Do, Transfer learning methods for low-resource speech accent recognition: A case study on Vietnamese language, Engineering Applications of Artificial Intelligence, vol. 132, p. 107895, 2024. 10.1016/j.engappai.2024.107895

[16] N. Lashkarashvili, W. Wu, G. Sun, and P. C. Woodland, Parameter Efficient Finetuning for Speech Emotion Recognition and Domain Adaptation, ICASSP 2024 - 2024 IEEE International Conference on Acoustics, Speech and Signal Processing (ICASSP), 2024. 10.1109/ICASSP48485.2024.10446272

[17] D. Grangier, A. Katharopoulos, P. Ablin, and A. Hannun, Need a Small Specialized Language Model? Plan Early!, arXiv preprint 2402.01093, 2024. 10.48550/arXiv.2402.01093

[18] Y. Liu, S. Agarwal, and S. Venkataraman, AutoFreeze: Automatically Freezing Model Blocks to Accelerate Fine-tuning, arXiv preprint 2102.01386, 2021. https://arxiv.org/abs/2102.01386

[19] H. Liu, X. Liu, Q. Kong, W. Wang, and M. D. Plumbley, Learning Temporal Resolution in Spectrogram for Audio Classification, arXiv preprint 2210.01719v3, 2024. 10.48550/arXiv.2210.01719

[20] S. W. Fu, T. Y. Hu, Y. Tsao, and X. Lu, Complex spectrogram enhancement by convolutional neural network with multi-metrics learning, arXiv preprint 1704.08504, 2017. 10.48550/arXiv.1704.08504

[21] Q. Yu and R. Zhou, Amplitude and Phase Information Interaction for Speech Enhancement Method, Applied Sciences, vol. 13, no. 14, p. 8025, 2023. 10.3390/app13148025

[22] A. Vaswani, N. Shazeer, N. Parmar, et al., Attention is All You Need, NeurIPS 2017. [Online] Available: https://arxiv.org/abs/1706.03762.

[23] M. A. Shah, D. Solans, M. A. Heikkilä, B. Raj, and N. Kourtellis, Speech Robust Bench: A Robustness Benchmark for Speech Recognition, arXiv preprint 2403.07937v2, 2024. https://arxiv.org/abs/2403.07937

[24] K. Papineni, S. Roukos, T. Ward, and W.-J. Zhu, BLEU: A method for automatic evaluation of machine translation, Proceedings of the 40th Annual Meeting of the Association for Computational Linguistics (ACL 2002).

[25] Promila Ghosh, MediBeng (Revision b05b594), 2025. [Online]. Available: https://huggingface.co/datasets/pr0mila-gh0sh/MediBeng. DOI: 10.57967/hf/5187. [Accessed: Apr. 22, 2025].

[26] Hugging Face, Hugging Face: The AI community building the future, [Online]. Available: https://huggingface.co. [Accessed: Apr. 22, 2025].

[27] OpenAI, GPT: Generative Pre-trained Transformer for Natural Language Processing, [Online]. Available: https://openai.com/gpt. [Accessed: Apr. 22, 2025].

